# Performance of Cardiovascular Polygenic Risk Scores in Carotid Stenosis Identification

**DOI:** 10.64898/2026.06.22.26356289

**Authors:** Tiffany R. Bellomo, Anika Misra, Tianrun Cai, Lucia Sanchez Garcia, Aniruddh P. Patel, Alyssa M. Flores, Taylor Nordan, Tetsushi Nakao, Zhi Yu, Matthew J. Eagleton, Akl C. Fahed, Pradeep Natarajan

## Abstract

**Background:** Clinically significant carotid stenosis remains a major cause of ischemic stroke (IS), yet prediction of disease progression is limited. Polygenic risk scores (PRSs) for coronary artery disease (CAD) and peripheral artery disease (PAD) have demonstrated associations with atherosclerosis burden and major cardiovascular disease (CVD) events, but whether these insights extend to carotid stenosis is unclear. We evaluated the association and discriminative performance of validated PRSs for CAD, PAD, IS, and carotid intima-media thickness (cIMT) with carotid stenosis among participants of the Mass General Brigham Biobank (MGBB).

**Methods:** Carotid stenosis was identified in genotyped MGBB participants using validated ICD-and CPT-based phenotyping algorithms. Logistic regression adjusted for age, sex, and 10 ancestry principal components assessed PRS associations. Incremental discrimination was evaluated using changes in Harrell’s C-statistic.

**Results:** Compared with 52,636 controls, 670 participants with carotid stenosis were more frequently male (61.5% vs 44.1%), older (70.8 SD 9.0 vs 53.4 SD 17.2 years), and more likely to be European (95.7% vs 84.1%). The IS (OR 1.31, 95% CI 1.21–1.41), CAD (OR 1.62, 95% CI 1.50–1.75), and PAD (OR 1.66, 95% CI 1.54–1.80) PRSs were each associated with carotid stenosis (all p<0.0001), while the cIMT PRS was not (OR 1.04, 95% CI 0.97–1.13; p=0.28). The PAD PRS demonstrated the greatest improvement in discrimination beyond age, sex, and ancestry (ΔC-statistic 0.017; C-statistic 0.845, 95% CI 0.833–0.856). A fully adjusted model incorporating established CVD risk factors achieved a C-statistic of 0.852 (95% CI 0.841–0.862), with modest further improvement after PAD PRS inclusion (ΔC-statistic 0.008). Individuals in the top 5% of the PAD PRS distribution and top 4% of CAD PRS demonstrated 3-fold greater odds of carotid stenosis.

**Conclusions:** A PAD and CAD PRS may help identify individuals at high likelihood for carotid stenosis, though broad discriminative performance remains limited. These findings support further investigation of CVD PRSs as adjunctive risk stratification tools.

**Clinical Perspective:** *What’s New?:*

- Polygenic risk scores (PRSs) for peripheral artery disease (PAD), coronary artery disease (CAD), and ischemic stroke (IS), and carotid intima media thickness (cIMT) were each independently associated with carotid stenosis presence.
- The PAD PRS demonstrated the strongest discrimination for carotid stenosis, whereas the ischemic stroke PRS demonstrated the weakest and the cIMT PRS demonstrated no discriminatory ability.
- Both the PAD and CAD PRS improved discrimination beyond traditional cardiovascular risk factors and identified carotid stenosis without IS, with IS, and overall, with up to 3-fold greater odds of carotid stenosis.

*Clinical Implications:*

- The PAD and CAD PRS may serve as an adjunctive tool for carotid stenosis identification beyond traditional cardiovascular risk factors.
- Development and validation of carotid-specific PRSs in diverse populations may further improve precision risk assessment for carotid atherosclerotic disease.

## INTRODUCTION

Stroke remains a leading cause of morbidity, mortality, and long-term disability worldwide^1^. Approximately 70% of all strokes are ischemic in nature, and up to 20% are attributable to cervical internal carotid artery stenosis^2^. Carotid stenosis often remains asymptomatic for extended periods before progressing to morbid complications, and surgical intervention is highly effective at preventing stroke in appropriately selected patients^3^. However, the inability to reliably predict which patients will progress from asymptomatic to symptomatic disease represents a critical clinical challenge^4–6^, limiting accurate identification of those who would derive the greatest benefit from preventive surgical intervention^7^.

Human genetics has been successfully leveraged to gain insight into the pathophysiology of atherosclerosis and disease progression. The genetic architecture of atherosclerotic diseases in the population is largely polygenic, with many loci exhibiting pleiotropic effects across multiple cardiovascular disease (CVD) risk factors^8,9^. Polygenic risk scores (PRSs), which aggregate the effects of common variants identified through genome-wide association studies (GWAS), quantify inherited susceptibility to atherosclerotic disease and may identify high-risk individuals well before traditional risk factors become clinically informative, thereby guiding earlier preventive interventions^10,11^. Validated PRSs have been developed for coronary artery disease (CAD)^12^, peripheral artery disease (PAD)^13^, ischemic stroke (IS)^14^, and carotid intima-media thickness (cIMT)^15^, and in conjunction with traditional clinical risk factors, these scores have demonstrated the ability to stratify risk across healthy populations and are already being applied clinically^16^. Generally, CAD and PAD PRS associate with overall subclinical atherosclerosis burden as well as high risk features^17,18^.

Despite advances in polygenic risk stratification for other atherosclerotic vascular diseases^13,19^, the extent to which these findings relate to carotid stenosis, an important risk factor and cause of ischemic stroke (IS) remains unknown. Prior genetic studies of carotid disease have been largely restricted to cIMT ^20,21^, a research measurement not routinely used to trend disease progression in clinical practice^22^ and not robustly associated with incident stroke^23,24^. Furthermore, the low prevalence of clinically ascertained carotid stenosis in population-based cohorts constrains the sample sizes necessary to develop a carotid-specific PRS, prompting investigation into whether PRSs for other atherosclerotic CVDs may serve as informative predictive tools in this population.

The Mass General Brigham Biobank (MGBB) is an ongoing hospital-based cohort study enriched with longitudinal electronic medical record (EHR), genomic, and health data^25^. In this study, we sought to evaluate the performance of validated CVD PRS developed for CAD, PAD, IS, and cIMT, in patients with carotid stenosis to determine their value for risk stratification beyond traditional cardiovascular risk factors.

## METHODS

### Design and Study Population

MGBB is a hospital-based cohort study that began enrollment in 2010 within the Mass General Brigham health system, the largest academic healthcare network in Massachusetts^26^. EHR data was queried through the Mass General Brigham Research Patient Data Registry (RPDR)^27^. The RPDR was queried for all patients with available data from January 2010 through January 2025. This study was approved by the Massachusetts General Brigham Institutional Review Board in accordance with the Declaration of Helsinki under IRB #2021P002228. The need for informed consent was waived given the retrospective nature of this study.

At the time of this analysis, more than 140,000 participants had been enrolled, of whom 53,306 had available genotyping data linked to the EHR, as previously described^28^. Participants were genotyped using the Illumina Global Screening Array. Quality control measures included removal of insertion-deletion variants, multiallelic sites, variants with missingness exceeding 2%, and variants with a minor allele count less than 3. Genotype imputation was performed using the TOPMed r2 multi-ancestry reference panel via the TOPMed Imputation Server^29^. Ancestry inference was performed using principal component (PC) projection based on the 1000 Genomes Project reference data, with genetic similarity groups assigned using k-nearest neighbor (kNN) classification.

### PRS Construction

All training data for each PRS were external to MGBB. The PRS for IS in this analysis was derived from the integrative PRS model published by Mishra et al. (PGS002724), described in detail in the GIGASTROKE consortium^14^ (Supplemental Table 1). Briefly, this PRS was constructed by combining GWAS summary statistics from multiple stroke subtypes, including cardiometabolic and atherosclerotic, and vascular risk trait analyses using elastic-net logistic regression, distinguishing it from single-trait polygenic scores. For the European ancestry model, training was performed in 1,003 prevalent IS cases and 8,997 controls from the Estonian Biobank, incorporating 10 GIGASTROKE GWAS analyses across stroke subtypes and 12 vascular risk trait GWAS analyses. In the prior work, model performance was evaluated on an independent, non-overlapping sample of 102,099 participants, achieving a ΔC-index of 0.027 over a base model adjusted for age, sex, and the top five principal components of ancestry.

The PRS for CAD in this analysis was derived from GPSMult (PGS003725), a validated multi-ancestry, multi-trait genomic risk score previously described in detail^12^. Briefly, GPSMult was constructed using a two-layer framework in which ancestry-specific CAD PRS were first generated from summary statistics drawn from multiple large-scale GWAS conducted in diverse populations-including FinnGen, the Million Veteran Program, Biobank Japan, and CARDIOGRAMplusC4D-using the LDpred2 Bayesian shrinkage method applied to HapMap3 variants. These ancestry-specific scores were linearly combined into a single multi-ancestry CAD score (layer 1), which was then further combined with analogously constructed polygenic scores for CAD-related traits, including LDL cholesterol, HDL cholesterol, triglycerides, type 2 diabetes, systolic blood pressure, and body mass index, to generate the final GPSMult (layer 2). Mixing weights at each layer were estimated via logistic regression with feature selection guided by the Akaike information criterion. The final score was validated in independent cohorts across multiple ancestries within the UK Biobank, the Million Veteran Program, and the Genes & Health study.

The PRS for PAD was derived from GPSPAD (PGS005217), a validated multi-ancestry, multi-trait genomic risk score previously described in detail by Flores AM et al.^13^, constructed using the same two-layer framework as GPSMult. Briefly, ancestry-specific PAD polygenic scores were generated from GWAS summary statistics across five ancestral groups-African, East Asian, European, Latino, and South Asian-using LDpred2-auto applied to HapMap3 variants with ancestry-specific 1000 Genomes linkage disequilibrium reference panels. Feature selection and mixing weight estimation at both layers were performed using LASSO logistic regression, resulting in 54 ancestry-specific scores across 11 traits contributing to the final GPSPAD. The score was trained and validated within the UK Biobank and externally validated in the All of Us Research Program.

The PRS for cIMT was derived from cIMT PRS (PGS002184), a validated multi-ancestry, multi-trait genomic risk score previously described by Privé F. et al^15^. Briefly, 245 PRS were generated from individual-level UK Biobank genotype data using penalized regression (lasso) and LDpred2-auto applied to HapMap3 variants, with ancestry-specific linkage disequilibrium reference panels derived from the UK Biobank. cIMT was defined by UK Biobank data fields capturing mean cIMT at 120°, 150°, 210°, and 240° (fields 22671, 22674, 22677, and 22680, respectively). Scores were trained in participants of Northwestern European ancestry (n=391,124) and assessed for portability across eight additional ancestry groups within the UK Biobank; external validation was not performed.

Within the MGBB cohort used in this study, computed PRS were residualized for the first ten principal components of genetic ancestry and scaled to a mean of 0 and standard deviation of 1 prior to analysis.

### Outcome Definitions

Carotid stenosis outcome definitions included carotid stenosis total, carotid stenosis only, and carotid stenosis with IS. Carotid stenosis total was defined based on ICD and CPT codes for carotid stenosis and revascularization at the time of enrollment in MGBB, validated with manual chart adjudication with a positive predictive value at around 90% for greater than 20% stenosis (Supplemental Table 2). Carotid stenosis total was further stratified based on the presence or absence of IS at the first occurrence of a qualifying event identified using the ICD- and CPT-based algorithm (Supplemental Table 3), resulting in the carotid stenosis with IS and carotid stenosis only groups, respectively. The algorithm for IS was developed by the MGBB where expert physicians had individual-level data access for adjudication of outcomes^30^.

### Demographics and Comorbidities

Analyses were performed using both demographic and common comorbidities related to carotid stenosis^3^. Age was defined at enrollment in MGBB, and sex was recorded as female or male. Current smoking was defined as lifetime smoking of at least 100 cigarettes and currently without cessation. Relevant comorbidities included diabetes, hypertension, hyperlipidemia, chronic kidney disease (CKD), and lipid levels were ascertained at the time of enrollment. Diabetes was defined as glycated A1c ≥6.5% or physician diagnosis. Hypertension was defined as a diagnostic code for systemic hypertension or prescription of an antihypertensive medication. Hyperlipidemia was defined as total cholesterol ≥200 mg/dL or statin prescription. CKD status was determined based on estimated glomerular filtration rate calculated using the CKD Epidemiology Collaboration equation with a cutoff of less than 60. Relevant medications included statin, antiplatelet, and anticoagulation medications. Antiplatelet medications were defined as aspirin, clopidogrel, prasugrel, dipyridamole, or ticagrelor. Anticoagulant medications were defined as taking a direct oral anticoagulant, low molecular weight heparin, or warfarin.

### Statistical Analysis

Descriptive statistics were calculated for all covariates stratified by carotid stenosis status. Continuous variables are reported as median [IQR] and compared using the Mann-Whitney U test; categorical variables are reported as proportions with counts and compared using the chi-square test, as displayed in Table 1. Logistic regression models adjusted for age, sex, and the first 10 PCs of genetic ancestry were used to evaluate the association of established CVD risk factors and each PRS with the outcome of carotid stenosis. Carotid stenosis cases were further stratified into those with and without subsequent IS to assess whether associations differed by clinical severity. The discriminative performance of each model was quantified using Harrell’s C-statistic. All associated CVD risk factors and PRS were used to construct a second set of models, in which each PRS was added individually to a model comprised of age, sex, the first 10 principal components of genetic ancestry, and the following traditional CVD risk factors: diabetes, hyperlipidemia, hypertension, chronic kidney disease, current smoker, statin prescription, antiplatelet prescription, anticoagulant prescription. The incremental improvement in discrimination was assessed as the ΔC-statistic relative to the base model. Finally, PRS threshold values corresponding to 2-fold, 2.5-fold, and 3-fold increases in odds of carotid stenosis relative to the population median were determined for each score. In all analyses, a two-sided p-value less than 0.05 and a 95% confidence interval excluding unity were considered evidence of statistical significance. All statistical analyses were performed in R (version 4.4.1).

**Table 1.** Summary of baseline characteristics of individuals studied from the MGBB by carotid stenosis. P-values are reported based on Chi-square tests for categorical variables and Wilcoxon rank-sum tests for continuous variables. Abbreviations: SD, Standard Deviation; IQR, Interquartile Range; MGBB, Mass General Brigham Biobank.

| Variable | Carotid Stenosis<br>Absent<br>(n=52,636) | Carotid Stenosis<br>Present<br>(n=670) | P-Value |
| --- | --- | --- | --- |
| Age (mean (SD)) | 53.4 (17.2) | 70.8 (9.0) | <0.0001 |
| Male Sex (%) | 23,214 (44.1%) | 412 (61.5%) | <0.0001 |
| Genetic Ancestry (%) |  |  | <0.0001 |
| African | 2,882 (5.5%) | 10 (1.5%) |  |
| Ad Mixed American | 3,835 (7.3%) | 16 (2.4%) |  |
| Eastern Asian | 1,064 (2.0%) | 0 (0.0%) |  |
| European | 44,251 (84.1%) | 641 (95.7%) |  |
| South Asian | 604 (1.1%) | 3 (0.4%) |  |
| Diabetes (%) | 4,628 (9.8%) | 219 (32.7%) | <0.0001 |
| Hyperlipidemia (%) | 25,283 (48.0%) | 532 (79.4%) | <0.0001 |
| Hypertension (%) | 36,883 (70.3%) | 664 (99.3%) | <0.0001 |
| Chronic Kidney Disease (%) | 6,571 (16.1%) | 260 (44.2%) | <0.0001 |
| Coronary Artery Disease (%) | 294 (43.9%) | 3,417 (7.2%) | <0.0001 |
| Peripheral Arterial Disease (%) | 1,565 (3.3%) | 278 (41.6%) | <0.0001 |
| Ischemic Stroke (%) | 854 (1.6%) | 112 (16.7%) | <0.0001 |
| Current Smoker (%) | 8,442 (17.7%) | 130 (20.1%) | 0.1237 |
| Statin Prescription (%) | 18,758 (35.6%) | 522 (77.9%) | <0.0001 |
| Antiplatelet Prescription (%) | 20,040 (38.1%) | 524 (78.2%) | <0.0001 |
| Anticoagulant Prescription (%) | 6,363 (12.1%) | 220 (32.8%) | <0.0001 |

## RESULTS

### Baseline Characteristics

A total of 670 cases and 52,636 control participants with genetic profiles were included in this analysis (Table 1). Compared with controls, participants with carotid stenosis were more frequently male (61.5% vs 44.1%, p<0.0001) and older with a mean age of 70.8 (SD 9.0) compared to 53.4 (SD 17.2). Ancestry composition differed significantly between groups, with a higher proportion of European ancestry among patients with carotid stenosis (95.7% vs 84.1%, p<0.0001). Participants with carotid stenosis had substantially higher rates of clinical CVD risk factors, including diabetes (32.7% vs 9.8%), hyperlipidemia (79.4% vs 48.0%), hypertension (99.3% vs 70.3%), and CKD (44.2% vs 16.1%). Patients with carotid stenosis were also more likely to be prescribed statins (77.9% vs 35.6%), antiplatelet therapy (78.2% vs 38.1%), and anticoagulants (32.8% vs 12.1%) (all p<0.0001). Current smoking status did not differ significantly between groups (20.1% vs 17.7%, p=0.12). Among those with carotid stenosis, 112 had an IS event after their carotid stenosis diagnosis (Supplemental Table 4).

### PRS and Risk Factor Associations

After adjusting for age, sex, and the first 10 PCs of genetic ancestry, three PRSs were significantly associated with total carotid stenosis per SD increase: the IS PRS (OR 1.31, 95% CI 1.21-1.41, p<0.0001), the CAD PRS (OR 1.62, 95% CI 1.50-1.75, p<0.0001), and the PAD PRS (OR 1.66, 95% CI 1.54-1.80, p<0.0001) (Figure 1). cIMT PRS was not significantly associated with carotid stenosis (OR 1.04, 95% CI 0.97-1.13, p=0.28). These associations were consistent across carotid stenosis total, carotid stenosis only, and carotid stenosis with IS. All established CVD risk factors were also positively associated with carotid stenosis (p<0.0001). Prescriptions for statins (OR 2.98, 95% CI 2.47-3.60, p<0.0001), antiplatelet agents (OR 2.83, 95% CI 2.34-3.42, p<0.0001), and anticoagulation (OR 1.81, 95% CI 1.53-2.15, p<0.0001) were positively associated with carotid stenosis, likely reflecting indication bias rather than causal association. When analyses were restricted to patients with carotid stenosis to evaluate predictors of subsequent IS, neither the PRSs nor clinical CVD risk factors were significantly associated with IS among those with carotid stenosis in the present sample size (all p>0.100) (Supplemental Figure 1). While we cannot rule out the possibility of reduced statistical power, as point estimates were all generally close to 1 for PRSs and clinical CVD risk factors.

**Figure 1.**
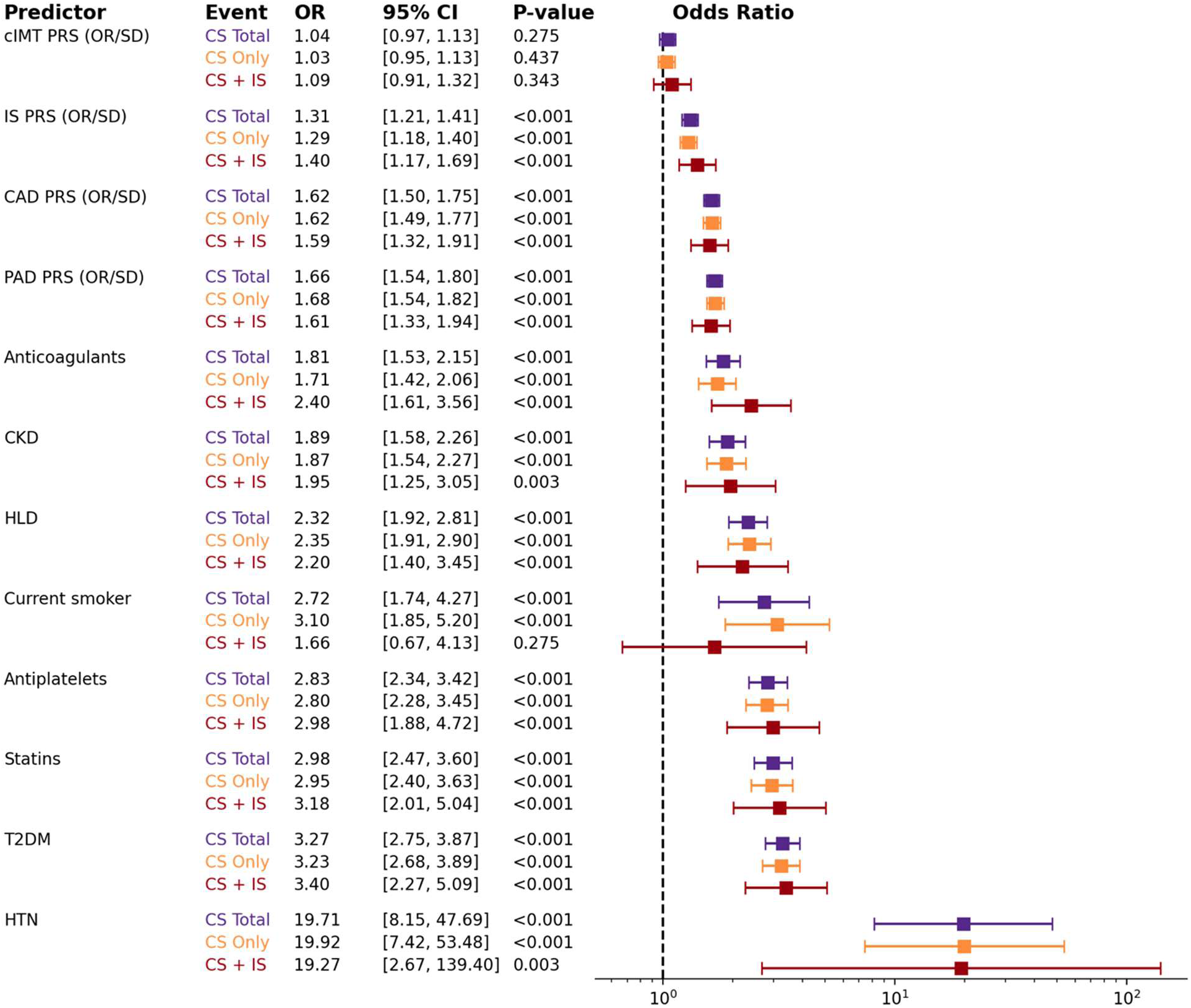
Association of cardiovascular polygenic risk scores and established atherosclerotic risk factors with carotid stenosis. OR with 95% confidence intervals are presented for three carotid stenosis phenotypes: total carotid stenosis (purple), carotid stenosis without ischemic stroke (orange), and carotid stenosis with subsequent ischemic stroke (dark red). All models were fit using logistic regression adjusted for age, sex, and the first 10 principal components of genetic ancestry. PRS are displayed as OR per standard deviation (SD). The dashed vertical line represents an OR of 1.0 (null effect). P-values were derived from two-sided t-tests implemented via the GLM function in R. PRS, polygenic risk score; CAD, coronary artery disease; PAD, peripheral artery disease; cIMT, carotid intima-media thickness; IS, ischemic stroke; CKD, chronic kidney disease; HLD, hyperlipidemia; T2DM, type 2 diabetes mellitus; OR, Odds ratios.

When comparing the change in C-statistic from a base model of age, sex, and the first 10 PCs of ancestry (C-statistic 0.827, 95% CI 0.815-0.839), the largest improvement was observed with the PAD PRS (ΔC-statistic 0.017; C-statistic 0.845, 95% CI 0.833-0.856), followed by the CAD PRS (ΔC-statistic 0.016; C-statistic 0.843, 95% CI 0.831-0.854), and the IS PRS (ΔC-statistic 0.006; C-statistic 0.833, 95% CI 0.821-0.845) (Figure 2, Supplemental Figure 2). A similar pattern was observed across carotid stenosis total and carotid stenosis only, with the exception of the carotid stenosis with IS group, in which CKD (ΔC-statistic 0.010; C-statistic 0.816, 95% CI 0.786-0.845), current smoking (ΔC-statistic 0.010; C-statistic 0.815, 95% CI 0.786-0.845), and anticoagulation use (ΔC-statistic 0.015; C-statistic 0.820, 95% CI 0.793-0.846) demonstrated relatively larger incremental improvements. When analyses were restricted to patients with carotid stenosis, CKD demonstrated the largest positive ΔC-statistic at 0.012 (C-statistic 0.608, 95% CI 0.549-0.666) (Supplemental Figure 3). However, all PRSs and CVD risk factors demonstrated poor overall discrimination when restricted to individuals with carotid stenosis (Supplemental Figure 4).

**Figure 2.**
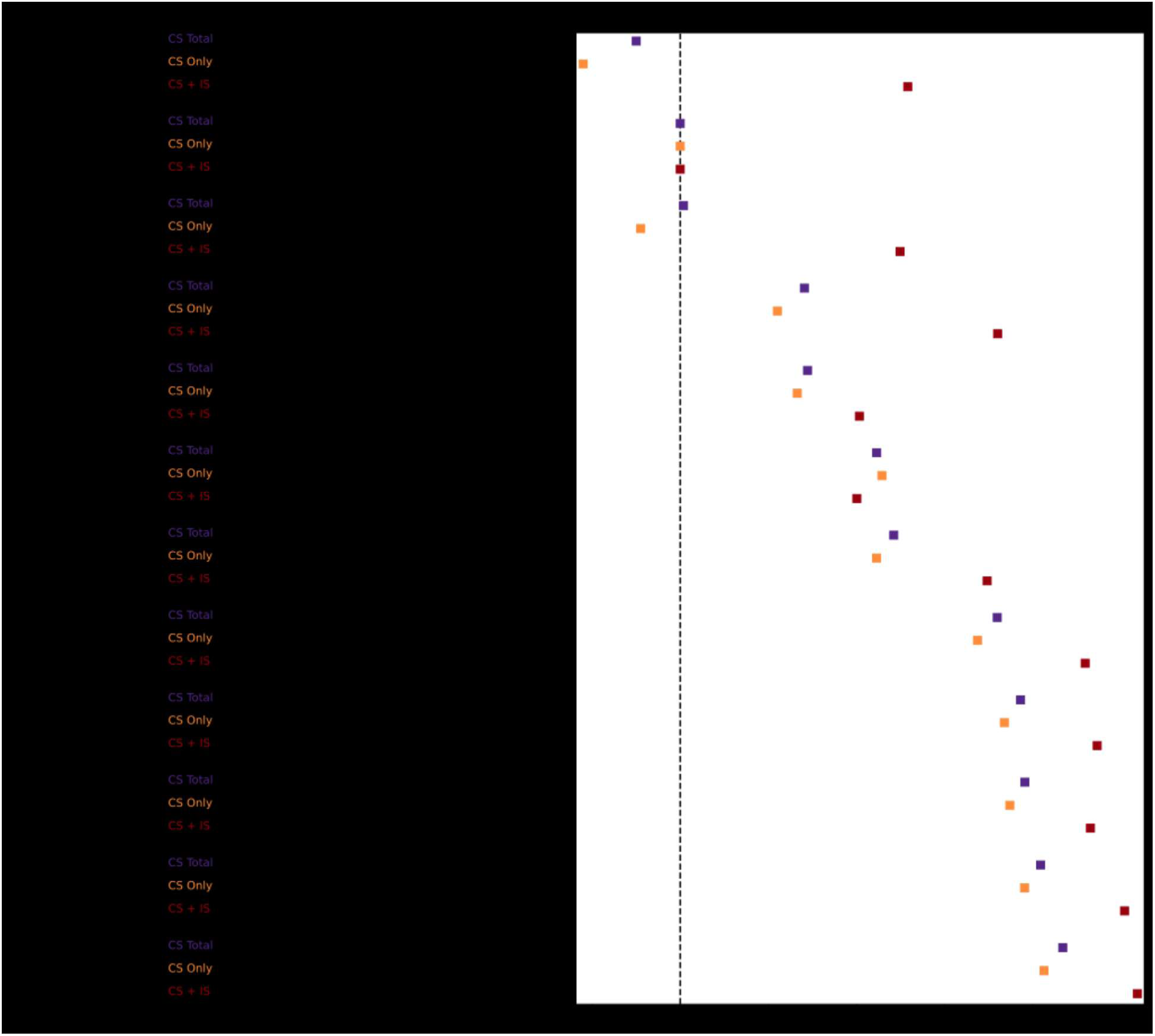
Incremental discriminative performance of cardiovascular polygenic risk scores and established atherosclerotic risk factors for carotid stenosis. The change in C-statistic (ΔC-statistic) is presented for each predictor across three carotid stenosis phenotypes: total carotid stenosis (purple), carotid stenosis without ischemic stroke (orange), and carotid stenosis with subsequent ischemic stroke (dark red). The ΔC-statistic reflects the incremental improvement in model discrimination relative to a base model of age, sex, and the first 10 principal components of genetic ancestry. Polygenic risk scores are evaluated per standard deviation (SD). The dashed vertical line represents a ΔC-statistic of 0 (no incremental improvement). All models were fit using logistic regression. PRS, polygenic risk score; CAD, coronary artery disease; PAD, peripheral artery disease; IS, ischemic stroke; CKD, chronic kidney disease; HLD, hyperlipidemia; T2DM, type 2 diabetes mellitus.

### Incremental Predictive Performance of PRS

A model incorporating all established CVD risk factors, including age, race, sex, the first 10 principal components of genetic ancestry, diabetes, hyperlipidemia, hypertension, chronic kidney disease, current smoker, statin prescription, antiplatelet prescription, anticoagulant prescription, demonstrated excellent discrimination for total carotid stenosis (C-statistic 0.852, 95% CI 0.841-0.862), carotid stenosis without IS (C-statistic 0.853, 95% CI 0.840-0.865), and carotid stenosis with subsequent IS (C-statistic 0.861, 95% CI 0.834-0.888) (Figure 3). The addition of each PRS to this model further improved discrimination, with the PAD PRS (ΔC-statistic 0.008; C-statistic 0.859, 95% CI 0.849-0.870) and the CAD PRS (ΔC-statistic 0.008; C-statistic 0.859, 95% CI 0.848-0.870) yielding the largest yet modest incremental improvement, followed by the IS PRS (ΔC-statistic 0.002; C-statistic 0.853, 95% CI 0.842-0.865). This pattern was consistent across carotid stenosis total, carotid stenosis only, carotid stenosis with IS. Among patients with established carotid stenosis, C-statistic values remained poor across all models, with the largest yet modest incremental contribution from the IS PRS being negligible (ΔC-statistic 0.001; C-statistic 0.642, 95% CI 0.589-0.694) (Supplemental Figure 5).

**Figure 3.**
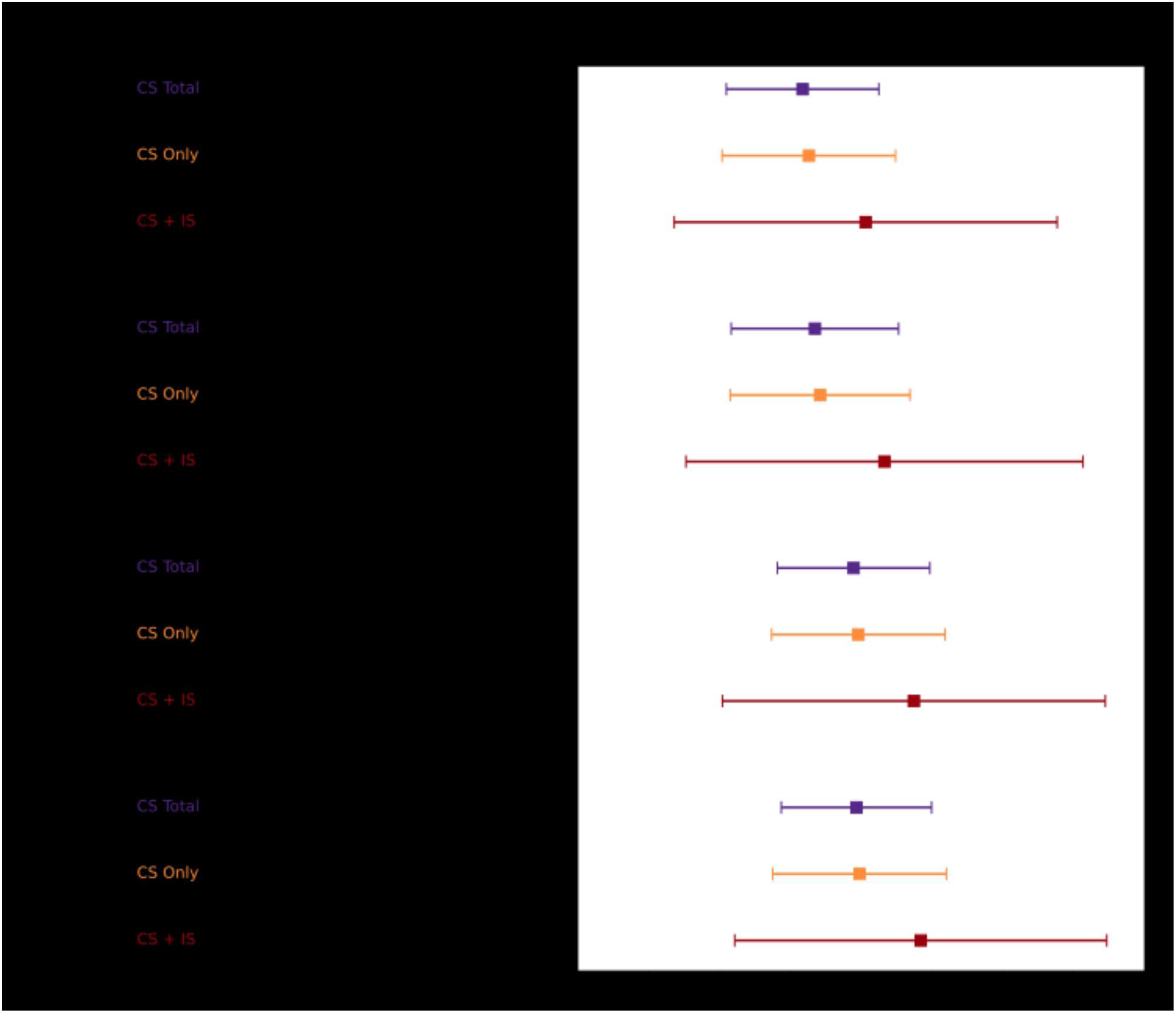
Absolute discriminative performance of cardiovascular polygenic risk scores added to a base model of traditional atherosclerotic risk factors for carotid stenosis. The C-statistic with 95% confidence intervals is presented for a base model of established cardiovascular risk factors alone and for models incorporating each PRS individually, across three carotid stenosis phenotypes: total carotid stenosis (purple), carotid stenosis without ischemic stroke (orange), and carotid stenosis with subsequent ischemic stroke (dark red). All models were fit using logistic regression adjusted for age, sex, and the first 10 principal components of genetic ancestry, diabetes, hyperlipidemia, hypertension, chronic kidney disease, current smoker, statin prescription, antiplatelet prescription, anticoagulant prescription. Polygenic risk scores are evaluated per standard deviation (SD). PRS, polygenic risk score; CAD, coronary artery disease; PAD, peripheral artery disease; IS, ischemic stroke; CKD, chronic kidney disease; HLD, hyperlipidemia; T2DM, type 2 diabetes mellitus.

### Population-Level PRS Risk Thresholds

Across all three PRSs and all carotid stenosis definitions, the association between risk score percentile and odds of carotid stenosis did not differ consistently by age at diagnosis (Supplemental Table 5). PRS-by-age interaction terms were non-significant for the PAD, CAD, and IS PRSs across all outcomes in both base and fully adjusted models (all p>0.05).

For the CAD PRS, the top 33%, 15%, and 4% of the population corresponded to 2-fold, 2.5-fold, and 3-fold greater odds of carotid stenosis relative to the population median, respectively (Table 2, Figure 4). Similarly, for the PAD PRS, the top 14%, 9%, and 5% of the population corresponded to 2-fold, 2.5-fold, and 3-fold greater odds of carotid stenosis, respectively. In contrast, for the IS PRS, only the top 3% of the population reached a 2-fold increase in odds. The performance was similar across carotid stenosis total, carotid stenosis only, and carotid stenosis with IS controls without disease (Supplemental Figure 6).

**Table 2.** Proportion of the MGBB population exceeding 2-fold, 2.5-fold, and 3-fold increased odds of carotid stenosis by polygenic risk score. Odds ratios (OR) with 95% confidence intervals and corresponding p-values are presented for each PRS threshold relative to the population median. Top percentile denotes the minimum PRS percentile at which the specified fold-risk threshold is achieved. Percent of the population reflects the proportion of MGBB participants exceeding each threshold. All models were fit using logistic regression adjusted for age, sex, and the first 10 principal components of genetic ancestry. PRS, polygenic risk score; CAD, coronary artery disease; CI, confidence interval; MGBB, Mass General Brigham Biobank.

| PRS | Fold Change | Percentile (%) | Cases (n) | Controls (n) | OR (95% CI) | P-Value | Population (%) |
| --- | --- | --- | --- | --- | --- | --- | --- |
| IS (PGS002724) | 2 | 98 | 35 | 1,564 | 2.07 (1.40-3.04) | 2.34E-04 | 3 |
| CAD (PGS003725) | 2 | 68 | 350 | 17,231 | 2.03 (1.63-2.54) | 3.45E-10 | 33 |
| CAD (PGS003725) | 2.5 | 86 | 195 | 7,797 | 2.56 (2.01-3.27) | 2.74E-14 | 15 |
| CAD (PGS003725) | 3 | 97 | 63 | 2,068 | 3.22 (2.33-4.46) | 1.51E-12 | 4 |
| PAD (PGS005217) | 2 | 87 | 181 | 7,278 | 2.06 (1.63-2.59) | 1.01E-09 | 14 |
| PAD (PGS005217) | 2.5 | 92 | 136 | 4,659 | 2.51 (1.95-3.21) | 4.84E-13 | 9 |
| PAD (PGS005217) | 3 | 96 | 87 | 2,577 | 3.03 (2.28-4.02) | 2.13E-14 | 5 |

**Figure 4.**
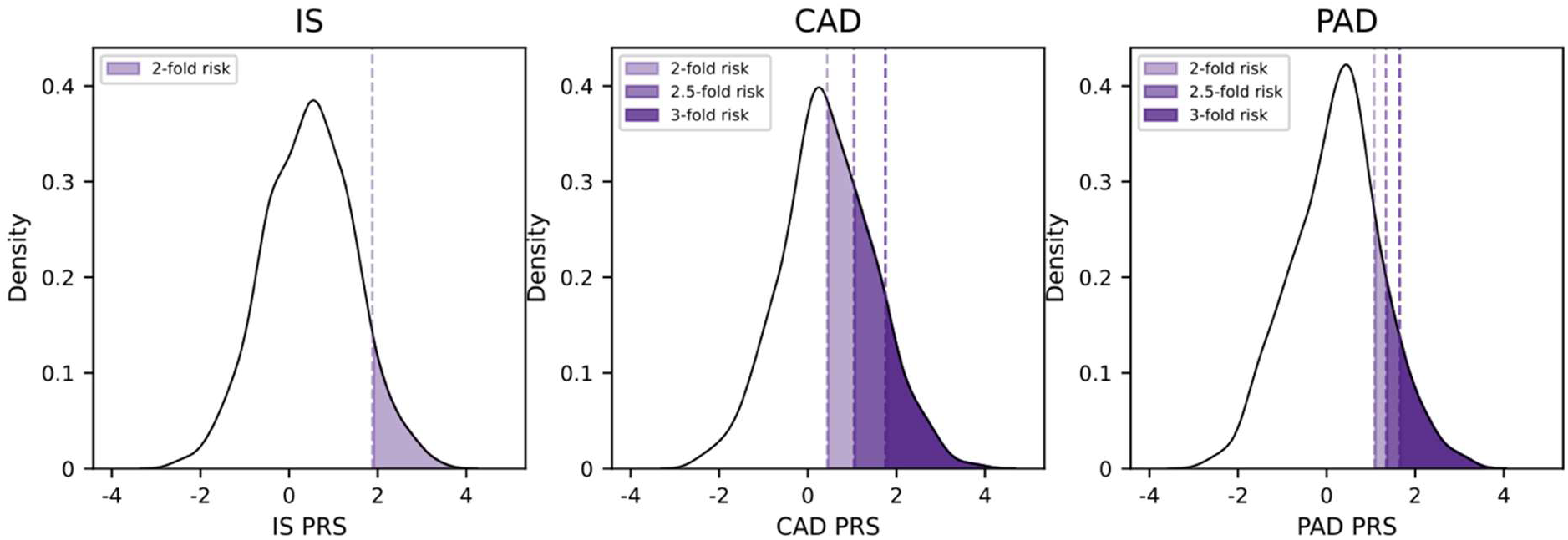
Proportion of the MGBB population exceeding 2-fold, 2.5-fold, and 3-fold increased odds of carotid stenosis by polygenic risk score. Panels display the population distribution of each PRS with shaded regions denoting the proportion of the MGBB population corresponding to 2-fold (light purple), 2.5-fold (medium purple), and 3-fold (dark purple) greater odds of carotid stenosis relative to the population median for: A) ischemic stroke PRS (PGS002724), B) CAD PRS (PGS003725), and C) PAD PRS (PGS005217). Dashed vertical lines denote the PRS thresholds corresponding to each risk level. Odds ratios were estimated using logistic regression adjusted for age, sex, and the first 10 principal components of genetic ancestry. PRS, polygenic risk score; CAD, coronary artery disease; PAD, peripheral artery disease.

## DISCUSSION

In this study, we demonstrate that PRSs for IS, CAD, and PAD were significantly associated with carotid stenosis. Although clinical risk factor models already demonstrated strong discrimination (C-statistic 0.852, 95% CI 0.841-0.862), the addition of the PAD PRS and the CAD PRS provided the largest further improvement (ΔC-statistic 0.008). Higher PAD PRSs were strongly associated with progressively greater odds of carotid stenosis, with the top 14%, 9%, and 5% of the population corresponding to 2-fold, 2.5-fold, and 3-fold greater odds of carotid stenosis, respectively. However, the IS PRS showed a substantially weaker association and did not reach a 3-fold risk threshold. Collectively, these findings suggest that carotid stenosis may share stronger genetic similarities with PAD than with IS, and that there may be value in incorporating PRSs into carotid stenosis identification.

Current efforts to identify patients with carotid stenosis at highest risk of stroke have centered on degree of luminal narrowing, with guidelines recommending carotid endarterectomy for asymptomatic stenosis exceeding 70%^3^, though the supporting evidence remains mixed and considerable variability persists in operative thresholds across clinical practice^4^. Beyond surgical candidacy, contemporary medical management targets traditional atherosclerotic CVD risk factors implicated in disease initiation and progression, including statin therapy^31^, antiplatelet agents^32^, and blood pressure control^33^. In this study, we demonstrate that a combination of traditional CVD risk factors yielded discrimination consistent with prior reports employing similar predictor sets^34^. However, optimized medical management of these risk factors does not uniformly prevent disease progression, and a meaningful proportion of patients continue to experience stroke despite guideline-directed therapy^35^. This residual risk suggests that conventional risk factors may not capture the heritable biological risk underlying carotid atherosclerosis.

Genetic risk stratification may improve identification of patients with clinically significant carotid stenosis. Prior studies evaluating clinical comorbidities and plaque characteristics associated with stroke risk, including hypertension^7^, ultrasound echolucency^36^, and intraplaque hemorrhage on MRI^37^, have identified important associations that have not translated into novel therapies or substantial changes in clinical practice. In this context, genetic studies may complement traditional CVD risk assessment by identifying individuals at elevated lifetime risk who could benefit from earlier screening, intensified surveillance, or preventive intervention^16^. However, prior genetic studies of carotid disease have largely focused on subclinical phenotypes such as cIMT^20,21^, a measurement not routinely used to monitor disease progression in clinical practice^22^, and one that has not been consistently associated with stroke^23,24^. Mendelian randomization studies have not supported a direct causal effect for cIMT on all stroke subtypes^38^. Although a PRS for cIMT has been developed^15^, it has not been externally validated and its utility in carotid stenosis specific phenotypes remains uncertain. In the present study, this cIMT PRS did not associate with carotid stenosis, further limiting its applicability.

Given PAD, CAD, and IS share many atherosclerotic risk factors and frequently co-occur^39,40^, we evaluated whether PRSs for these conditions could improve prediction of carotid stenosis. While one may have anticipated that the IS PRS would provide the greatest discriminative value given the established relationship between carotid stenosis and IS^41^, the IS PRS in this study demonstrated the weakest association among the three scores. This finding likely reflects the heterogeneous genetic architecture of ischemic stroke, which comprises multiple etiologically distinct subtypes whose combined genetic signal may dilute the specificity of a stroke PRS for carotid stenosis. Although the large artery stroke subtype most directly linked to carotid stenosis carries the highest heritability estimates among stroke subtypes^42^, the available stroke PRSs aggregate variants across all subtypes rather than isolating large artery disease specifically.

The strong epidemiologic co-occurrence of PAD and CAD with carotid stenosis provides biological rationale for the discriminative performance of their respective PRSs in the present study. The American Heart Association Scientific Statement on PAD health disparities reports that 50% of patients with PAD have concomitant CAD or cerebrovascular disease^43^. Further evidence includes large population based studies, where the Atherosclerosis Risk in Communities Study demonstrated that individuals with an ankle-brachial index below 0.90 had a significantly higher prevalence of preclinical carotid plaque^44^. Analysis of the Life Line Screening database demonstrated that patients with PAD had greater than 50% carotid stenosis prevalence of 19% compared with 3% among those without PAD, and multivariable regression identified PAD as an independent predictor of carotid stenosis^45^. A summative meta-analysis of over 4,000 patients with PAD estimated a higher prevalence, where around 25% had a greater than 50% carotid stenosis^46^. CAD also intersects with carotid stenosis at the clinical level, as myocardial ischemia from CAD remains the leading cause of perioperative complications following carotid artery surgery^47^.

In the present study, the PAD PRS demonstrated the highest predictive value for carotid stenosis, consistent with evidence showing that the genetic correlation between cIMT and PAD (rg = 0.45) is more than twice that observed between cIMT and CAD (rg = 0.21)^38^, suggesting that carotid atherosclerosis shares a greater proportion of its genetic architecture with PAD than with CAD^48^. Reflecting this relationship, the Society for Vascular Surgery guidelines recommend carotid screening in patients with lower extremity PAD, noting that the prevalence of ≥60% carotid stenosis in symptomatic PAD patients is likely more than 20%^43^. These findings are consistent with these recommendations. The present study also indicates that a PAD PRS may additionally have clinical utility in identifying individuals at elevated risk for carotid stenosis.

### Limitations

There are several limitations to the present study. First, MGBB is a volunteer-based cohort derived from a tertiary care center yet is enriched for clinically significant conditions. Second, our outcome definitions have inherent limitations, as they relied on diagnosis and procedure codes with some physician adjudication. Third, a significant proportion of PRSs in the PGS Catalog are derived from populations of primarily European genetic ancestry, limiting generalizability to more diverse populations and potentially exacerbating health disparities. Beyond PRS derivation concerns, the cohort itself may also have limited representation of non-European ancestry groups, further reducing external validity.

## CONCLUSIONS

A PAD and CAD PRS, but not a cIMT PRS, identified patients with clinically significant carotid stenosis, suggesting that genetic susceptibility to manifest atherosclerotic disease may be more informative than genetic susceptibility to subclinical carotid plaque burden. Future work is needed to determine whether a PAD PRS can improve outcomes related carotid atherosclerotic disease.

## Data Availability

These data can be shared for research upon request from the corresponding author, Dr. Pradeep Natarajan, MD, MMSc.

## ACKNOWLEDGEMENTS

The authors gratefully acknowledge the participants of the MGBB, whose voluntary contributions of data and biological samples form the foundation of this research. We also extend our appreciation to the MGBB staff for their ongoing efforts in data curation, participant engagement, and resource stewardship, which made this work possible.

## Abbreviations

PRS: Polygenic Risk Score
CS: Carotid Stenosis
PAD: Peripheral Artery Disease
CAD: Coronary Artery Disease
MGBB: Mass General Brigham Biobank
ICD: International Classification of Disease
SD: Standard Deviation
IQR: Interquartile Range
LDL: Low-density Lipoprotein
HDL: High-density Lipoprotein
HR: Hazard Ratio
CKD: Chronic Kidney Disease
cIMT: Carotid-intima Media Thickness

